# No evidence of interaction between *FADS2* genotype and breastfeeding on cognitive or other traits in the UK Biobank

**DOI:** 10.1101/2024.05.29.24308099

**Authors:** Giulio Centorame, Nicole M. Warrington, Gibran Hemani, Geng Wang, George Davey Smith, David M. Evans

**Author notes:** Correspondence: Giulio Centorame; Institute for Molecular Bioscience, 306 Carmody Road, St Lucia QLD 4072, Australia. Data share statement: the UK Biobank data described in the manuscript is available to all bona fide research upon application and approval. Code book and analytic code will be made publicly and freely available without restrictions at https://github.com/GiulioCentorame/FADS-by-breastfeeding/. List of abbreviations: AA, arachidonic acid; ALSPAC, Avon Longitudinal Study of Parents and Children; DBP, diastolic blood pressure; DHA, docosahexaenoic acid; EPA, eicosapentaneoic acid; E-risk, Environmental Risk Longitudinal Twin Study; GWAS, genome-wide association study; HDL, high-density lipoprotein; LA, linoleic acid; LC-PUFA, long-chain polyunsaturated fatty acids; LD, linkage disequilibrium; LDL, low-density lipoprotein; PC, principal component; PUFA, polyunsaturared fatty acids; SBP, systolic blood pressure; SES, socioeconomic status; SNP, single nucleotide polymorphism; TDI, Townsend deprivation index.

## Abstract

**Background:** Breastfeeding is hypothesised to benefit child health and cognitive functioning by providing long-chain polyunsaturated fatty acids (LC-PUFA), which are essential for brain development. In 2007, Caspi *et al.* found evidence in two cohorts for an interaction between genetic variation in the *FADS2* gene (a gene involved in fatty acid metabolism) and breast feeding on IQ. However, subsequent studies have provided mixed evidence for the existence of an interaction.

**Objective:** To investigate the relationship between genetic variation in the *FADS2* region, breastfeeding, and their interaction on traits putatively affected by their interplay in a large, population-based cohort with appropriate control for confounders in genetic associations.

**Methods:** We tested for the interaction in up to 335,650 individuals from the UK Biobank, over a range of cognitive functioning tests, as well as educational attainment and other traits thought to be influenced by breastfeeding, including cardiometabolic traits, reproductive success, and atopic allergy.

**Results:** *FADS2* alleles associated with an increase in docosahexaenoic acid (DHA) in blood serum (the C allele of rs174575) were associated with decreased verbal-numerical reasoning ( *p=* 2.28× 10^−5^) and triglycerides ( *p=* 1. 40× 10^−41^), increased reproductive success ( *p=* 3. 40× 10^−5^), total cholesterol ( *p=* 5. 28× 10^−36^), HDL ( *p=* 1. 42× 10^− 51^), and LDL cholesterol ( *p=* 1. 46× 10^− 21^). We observed no evidence of an interaction in any of the traits, regardless of the modelling strategy.

**Conclusions:** We failed to replicate any breastfeeding by genotype interactions on any cognitive or non-cognitive traits. We postulate that the previous positive findings are likely to be spurious, perhaps due to lack of appropriate control for latent population structure.

## Introduction

Breastfeeding is the recommended method of feeding infants for the first six months of their life (1,2). Over the last century, the scientific literature has reported evidence for the benefits of breastfeeding on infant mortality and health (3–7). An increasing body of evidence also suggests that breastfeeding may exert positive effects on cognitive outcomes, including observational studies that control for maternal intelligence and socioeconomic status (SES) (8,9), and a large randomised cluster trial (10).

One of the hypothesised mechanisms explaining the putative efficacy of breastfeeding on cognitive function, compared to other forms of infant feeding, is the presence of long-chain polyunsaturated fatty acids (LC-PUFA) in breastmilk. In particular, the presence of docosahexaenoic acid (DHA), an n-3 fatty acid (22:6(n-3)), and arachidonic acid (AA), an n-6 fatty acid (20:4(n-6)) (11,12), both of which were traditionally absent from commercial infant feeding formulas (13). DHA and AA are required for the development of the brain and retina (13,14) to synthesise membrane phospholipids such phosphoinositides, which regulate membrane proteins dedicated to transport and signalling (15), and phosphatidylserine, which have a structural role in cell membrane (16,17). They can be sourced through direct dietary intake of preformed acids or synthesised from precursors (e.g., from the conversion of linoleic acid [LA] and eicosapentaenoic acid [EPA], respectively) (18). It is generally understood that biosynthesis of DHA and AA in adult humans is limited (19,20)—except in women of childbearing age (20–25), while infants show a higher rate of bioconversion from exogenous sources of the precursors, LA and EPA (26,27). Breastmilk supplies infants with both the preformed DHA and AA and the precursors LA and EPA (28), that can subsequently be used by the infants themselves to biosynthesise DHA and AA.

Current evidence points to a beneficial effect of DHA and AA supplementation in standard infant feeding formulae for infant cognitive development, at a rate of around 1:1 (29,30), showing benefits on the development of executive functions (31–35) and activation and connectivity in cerebral cortex areas correlated with attention and cognition in term infants(36). However, trade regulations on their presence in standard infant feeding formulae are recent. Within the European Economic Area, regulations on commercially-available formulae permitted DHA supplementation in the 2000s (13,37), around the same time pre- and postnatal LC-PUFA supplementation became widespread (35), while the first regulations mandating set quantities were promulgated almost a decade later for DHA only (38,39), and even later for both (30). Until then, commercially-available formulae were prepared with ingredients containing negligible concentration of n-3 fatty acids (6,13,40).

DHA and AA biosynthesis are heavily influenced by *FADS2,* a gene on chromosome eleven that is a member of the fatty acid desaturase family. *FADS2* codifies enzymes present in the endoplasmic reticulum and mitochondria responsible the insertion of double bonds in fatty acid molecules, primarily on the 6^th^ carbon—carbon bond (Δ6 desaturase). This step is common to the metabolic pathway of both n-3 and n-6 fatty acids (41–43) and polymorphisms in *FADS2* associate with different levels of DHA and AA in serum and breastmilk (12,42,44–48).

In 2007, Caspi *et al.* (49) reported an interaction between a candidate intronic single nucleotide polymorphism (SNP) in the *FADS2* gene (rs174575), breastfeeding and childhood IQ. Participants who were breastfed and carried one or more copies of the C-allele (forward strand) putatively benefitted more from breastfeeding in terms of their IQ (with a 6.4 IQ points difference between breastfed and bottle-fed children) than GG homozygotes (who exhibited no significant difference in IQ between breastfed and bottle-fed infants). The study reported an interaction of similar form in two different cohorts: 1) the Dunedin Multidisciplinary Health and Development Study, a population-based cohort from New Zealand with child IQ measured at age 7, 9, 11, and 13, and 2) the Environmental Risk (E-risk) Longitudinal Twin Study (a twin-based cohort from the UK with IQ measured at age 5). Another SNP in the region, rs1535, (and in linkage disequilibrium [LD] with rs174575), displayed a similar interaction in the Dunedin cohort, but not in E-risk. The authors accounted for the potential confounding effects of age, sex, self-reported ancestry, and SES. Several subsequent studies have attempted to replicate Caspi *et al.* (49)’s findings, but have yielded mixed results (50–53).

The aim of the current study was to attempt to replicate the interaction between *FADS2* genotype, breastfeeding and IQ observed in Caspi *et al.* (49) in the UK Biobank (54). Our study benefits from the availability of genotype and phenotype data on over 335,000 individuals, as well as the presence of genome-wide SNP data, which together with appropriate statistical modelling, ensures the proper control of population stratification. We investigated the effect of *FADS* genotype and its purported interaction with breastfeeding using both a recessive coding at *FADS2* (i.e. used by many previous studies in this domain, including Caspi *et a*l. (49–51,53)) as well as an additive coding aligned to the DHA-increaser allele (since within locus additivity appears to be the rule rather than the exception in the case of common complex quantitative traits (55,56)). Finally, we leveraged the broad phenotyping present within the UK Biobank to investigate the association between *FADS2* variation and other traits that are purportedly influenced by LC-PUFA in breastmilk, including cardiometabolic phenotypes (12) and atopy (12,57–60).

## Methods

We used the STREGA checklist when writing our report (61).

### UK Biobank

UK Biobank is a large-scale volunteer-based prospective study containing health and lifestyle information on over 500,000 participants living in the United Kingdom, and is detailed elsewhere (54). Briefly, participants were recruited between 2006 and 2010 from a pool of 9.2 million individuals living in proximity to the assessment centres, where they provided baseline sociodemographic characteristics, reported on their health and lifestyle, and were assessed through questionnaires and physical measures. The participants were also asked to provide blood samples, which were processed for genotyping and metabolite blood assays, and asked to complete a follow-up questionnaire online (from 2014). A subset of participants (n = 20,319; https://biobank.ctsu.ox.ac.uk/crystal/field.cgi?id=110003) was followed up over three in-person assessment visits (first repeat assessment [2012-2013], imaging visit [2014-ongoing], first repeat imaging visit [2019-ongoing]).

Genotyping was performed using the UKBiLEVE array (n = 49,979) and UK Biobank AxiomArray (n = 488,377). To avoid possible genotyping errors, we excluded participants from our analyses with aneuploidy (n = 651), outliers for missingness and heterozygosity (n = 968), and genetic sex—reported sex mismatch (n = 378), as defined by UK Biobank (https://biobank.ctsu.ox.ac.uk/crystal/refer.cgi?id=531). We restricted our analyses to individuals who were unrelated to anyone else in the sample (up to the 3^rd^ degree) by excluding one individual for each kinship pair (n = 80,982), or that were excluded from the kinship estimation (n = 977), as identified by the UK Biobank using the KING analysis software (62) (described in Bycroft *et al.* (54)). We further excluded all the individuals that had no imputed genotypes available (n = 15,214) and participants who withdrawn their data from the study by the time of the analysis (N = 98). For our main analyses, we restricted participants to individuals of White British ancestry, as defined by the UK Biobank, using both self-reported information and genetic PCs (n = 335,650; https://biobank.ctsu.ox.ac.uk/crystal/field.cgi?id=22006; described in Bycroft *et al.* (54)).

This was done in order to account for differences in allele frequency across different ancestral populations and to match previous genetic studies of *FADS* and breast feeding, which have focused mainly on participants of European ancestry (49,50,53). We also included the first 40 genetic PCs, as provided by UK Biobank, in order to account for residual population stratification in the sample. Finally, we derived a subset of unrelated Non-White British individuals with data on breastfeeding (n = 69,858) to investigate the effect that uncorrected population stratification might have on the presence of genetic associations. A summary flowchart of the participant selection is presented in **Figure 1**.

**Figure 1:**
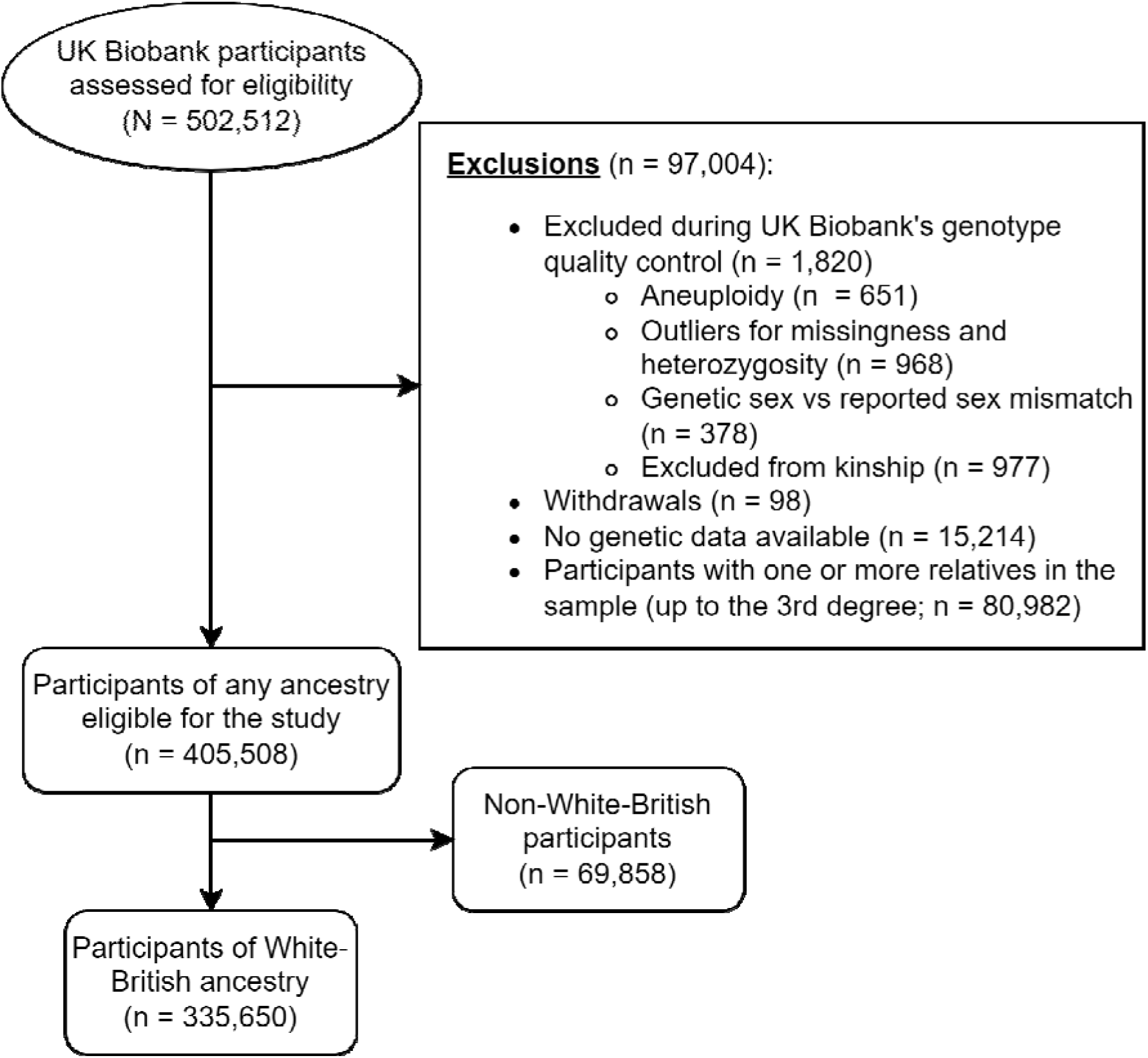
Participant selection flowchart. Overall sample size and exclusions are based on application 53641. Some participants fulfilled more than one exclusion criterion (e.g., excluded from kinship and outliers for missingness and heterozygosity).

#### Variant selection

We selected three coding variants examined in previous genetic association studies of *FADS2*, breastfeeding and IQ (49–53). Two of the variants (rs1535 and rs174583) were directly genotyped by both UKBiLEVE and the UK Biobank AxiomArray, while the third one (rs174575) was a high-quality imputed variant (IMPUTE4 INFO score > 0.99); all the variants followed Hardy-Weinberg Equilibrium (p < 0.05). Then, we calculated the LD across variants using LDBIRD (63). Summary information on the variants and their LD structure is available in **Supplementary Table 1**.

We extracted the allelic dosages (for rs174575) or direct genotype count (for rs1535 and rs174583) using plink 2.0 (64,65). We conducted statistical analyses assuming two different genetic effects on the outcomes:

- Additive allelic effect: we coded allelic dosages so that the effect allele increased levels of DHA in blood serum. Specifically, the A allele was the effect allele for rs1535, and the C allele was the effect allele for both rs174575 and rs174583.
- Recessive allelic effect (49,50,53): we modelled the SNP under the assumption of a recessive effect of the G-allele (for rs1535 and rs174575) or the T-allele (for rs174583), coded as 1 = homozygous for the recessive allele, 0 = carrier of the other allele. This is consistent with the coding of genotypes in most previous studies of *FADS2*, breastfeeding and IQ (49–51,53) (although it will most likely produce coefficients in the opposite direction to the additive coding). For the imputed variant (rs174575), we used the best-guess genotype to code according to a recessive model.

We determined the DHA-increasing allele by querying openGWAS (66) through the function *associations()* available in the R package *ieugwasr* (https://mrcieu.github.io/ieugwasr/), selecting data based on NMR metabolomics summary results from the UK Biobank (67).

Additionally, we investigated the association between the DHA-increasing alleles and n-3 fatty acid levels in blood serum, aligning the results to the DHA-increasing alleles. Due to the lack of data with comparable sample size for AA, we limited our analysis to LA (a precursor of AA) and n-6 fatty acids levels (**Supplementary Table 2**).

### Breastfeeding

UK Biobank participants were asked to report on whether they were breastfed or not as a baby at baseline and during follow-up visits. As the UK Biobank is skewed towards participants in late adulthood, we restricted our analyses only to the answers gathered during the initial assessment visit (2006–2010) to minimize recall errors. This led us to identify three strata: 1) participants who answered “Yes” (up to 180,484 participants), 2) participants who answered “No” (up to 73,981 participants), and 3) participants who answered “Do not know” and “Prefer not to answer” (labelled as “Missing”; up to 81,185 participants; baseline characteristics are presented in **Supplementary Table 3**).

### Outcomes

We investigated a range of outcomes, including cognitive measures, educational attainment, reproductive success, cardiometabolic traits, hayfever, rhinitis, and eczema. To attenuate the influence of outliers, we restricted our analysis only to participants with an outcome within 3 standard deviations of the mean. Sample size for the outcomes after exclusions and summary statistics for phenotypes and covariates are available in **Supplementary Tables 4 to 8**.

#### Cognitive measures

The participants were asked to complete a battery of questionnaires on cognitive performance and functioning at baseline, with repeated assessments during follow-up visits (including the imaging clinics; 2014-ongoing), and an online questionnaire in 2014. Some cognitive measures were introduced at different stages of assessment; for example, part of the battery of cognitive measures was administered only to a subset of individuals at baseline (e.g., numeric memory), while other measures (e.g., matrix pattern completion) were introduced only during the imaging follow-up visit and had not been assessed at baseline. Most notably, the questionnaire for verbal-numerical reasoning differed slightly between baseline and the online follow-up, due to the addition of one question. We natural log-transformed a subset of continuous traits that appeared to be positively skewed (viz., reaction time (68), visual memory (68,69), and trail-making paths 1 and 2 (70)), and then transformed the variables into z-scores. We added 1 point to the visual memory variable prior to the log-transformation in order to compensate for the extreme skewness on zero values (68), suggestive of floor effects on the measure.

When multiple data points for the cognitive measures were available, we took the first available instance, prioritising the assessment centre measure over the online measures wherever possible. Due to the different delivery mode across measures (and, in the case of the verbal-numerical reasoning scale, due to the different length of the questionnaire itself), all the models with outcomes that were administered both in the assessment centre and online (Verbal-numerical reasoning, Numeric memory, Visual memory, Trailmaking test, and Symbol digit substitution) were adjusted by the test delivery mode (0 = Assessment centre, 1 = Online).

We excluded participants who paused the assessment or who quit the assessment before completion, where information on the completion status was available.

#### Educational attainment

Educational attainment was operationalised as years of education required to achieve the highest qualification reported by the participant based on the ISCED 2011 equivalence to years of education (**Supplementary Table 9**). This operationalization has been used previously as a measure of educational attainment (71–74). We expect educational attainment to strongly correlate with cognitive development (75) and cognitive traits (76).

#### Reproductive success

We used number of offspring as a measure of reproductive success (77,78). We used reported number of children fathered for male participants and reported number of live births for female participants. We recoded values >10 to 10 to minimise the impact of outliers due to recall or data entry errors (79).

#### Cardiometabolic traits

We analysed blood assay measurements of total cholesterol, high density lipoprotein (HDL) cholesterol, low density lipoprotein (LDL) cholesterol, and triglycerides. Measures of systolic and diastolic blood pressure (SBP and DBP, respectively) were taken at baseline and follow up, with two readings for each assessment taken with a 1-minute interval. Readings were taken preferably with an automated machine and assessed manually only when the automated reading failed. We averaged the two measurements for each time point for both systolic and diastolic blood pressure. We selected the earliest available automatic measurement, where available; otherwise, we used the earliest available manual measurement. We accounted for blood pressure medication use by adding 15 mmHg to systolic blood pressure when the participants were under blood pressure lowering medications, and 10 mmHg for diastolic blood pressure (80). Body mass index (BMI; *kg*/*m*^2^) was calculated by UK Biobank from the height (m) and weight (kg) measures taken at baseline from the assessment centres.

We also investigated four cardiometabolic diseases using the self-reported diagnoses from the assessment centre’s touchscreen health questionnaires, including angina pectoris, myocardial infarction, stroke, and type-II diabetes. Participants were coded as type-II diabetes cases if they replied “Yes” to the question “Has a doctor ever told you that you have diabetes?” and “No” to “Did you start insulin within one year of your diagnosis of diabetes?” at any instance. The age covariate was set to the minimum age when they reported the diagnosis to the UK Biobank or the maximum age without reporting a diagnosis. Participants who responded “Prefer not to answer” were set to missing.

#### Hayfever, allergic rhinitis, or eczema

Finally, we investigated the relationship between *FADS2* and breastfeeding on hayfever, allergic rhinitis, or eczema from the assessment centre’s touchscreen questionnaire. We could not differentiate between the three conditions as no further question was asked of the UK Biobank participants to distinguish between them. To control for possible cohort effects in reporting atopic dermatitis, we used date of birth as a time covariate. Participants who responded “Prefer not to answer” or did not answer the question were set to missing.

### Statistical Modelling

To test for the association between breastfeeding and our outcomes, we fit:

1. A univariate model

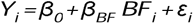

2. A model adjusting for the additive effect of age and sex:

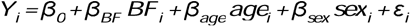

3. A model adjusting for the additive effect of age, sex, and Townsend deprivation index (TDI):

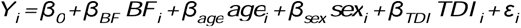

4. A model adjusting for the additive effect of age, sex, TDI, and 40 genetic PCs:

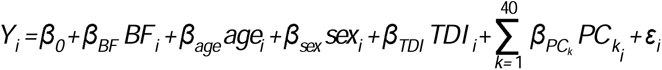

where for every participant *i*, *Y* is the outcome of interest, β_0_ is the intercept, *BF* is the participant’s breastfeeding status (where 0 = bottle-fed, 1 = breastfed), with effect β*_BF_* on the outcome, *age* is the participant’s age at assessment, with effect β*_age_*, *sex* is the participant’s sex (where 0 = female, 1 = male), with effect β*_sex_*, *TDI* is the participant’s TDI, with effect β*_TDI_*, *P_C_ki__* is the value of the *k* th genetic PC for participant *i*, with effect β*_PC_k__*, and ε*i* is a random error term.

To test for the *FADS*-by-breastfeeding interaction, we fit

1. An unadjusted interaction model:

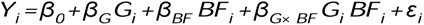

2. A model adjusting for the additive effect of age and sex:

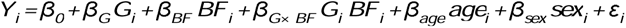

3. A model adjusting for the additive effect of age, sex, and 40 genetic PCs:

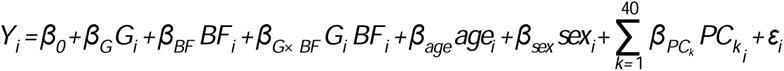

4. A model adjusting for the additive effect of age, sex, and PCs, and all the pairwise interactions across covariates (81):

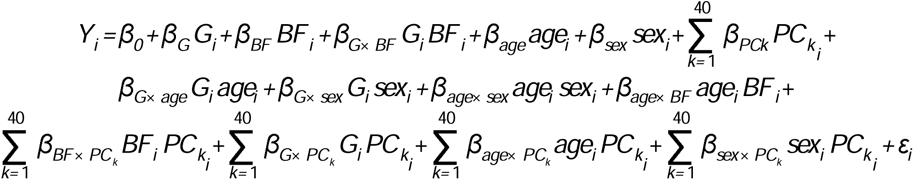

where *G* is the participant’s genotype, modelled using either a recessive coding (49,50,53) or an additive coding, counting the DHA-increasing allele (i.e., *A* for rs1535, *C* for rs174575, *C* for rs174583), with effect β*G* on the outcome, β*_G_*_× *BF*_ is the effect of the gene-by-environment interaction between genotype *G* and breastfeeding status *BF*, β*_G_*_× *age*_ is the effect of the interaction between *G* and *age*, β*G*× *sex* is the effect of the interaction between *G* and *sex* β*_age_*_× *sex*_ is the effect of the interaction between *age* and *sex*, β*_age_*_× *BF*_ is the effect of the interaction between *age* and *BF*, β*_sex_*_× *BF*_ is the effect of the interaction between *sex* and *BF*, β*_G_*_× *PC*_*_k_* is the effect of the interaction between *G* and the *k* th *PC*, β*_BF_*_× *PC*_*_k_* is the effect of the interaction between *BF* and the *k* th *PC*, β*_age_*_× *PC*_*_k_* is the effect of the interaction between *age* and the *k* th *PC*, and β*_sex_*_× *PC*_*_k_* is the effect of the interaction between *age* and the *k* th *PC*. To better visualize the results, we also fit the above models to each stratum of breast-feeding (breastfed, bottle-fed, or missing) without the interaction term.

To better visualize the results, we also fit the above models to each stratum of breast-feeding (breastfed, bottle-fed, or missing) without the interaction term.

For all analyses, we fit linear regression if the outcome was continuous, and logistic regression if the outcome was binary. Additionally, if the same cognitive measure was assessed both in person (at the assessment centre or during the imaging visits) and online, we added an additional covariate to all models to adjust for possible differences due to the delivery format and, in the case of verbal-numerical reasoning, a different number of questions in the questionnaire.

We restricted our analyses to participants with complete data on all variables in the given model.

## Results

### Phenotypic associations

Consistent with many previous studies (7,9), being breastfed as a baby was associated (p < 0.05) with improved cognitive function after controlling for covariates—particularly age and sex (**Figure 2** and **Supplementary Table 10 and 11**). This included tests for verbal-numerical reasoning, reaction time, numeric memory, paired associate learning, matrix pattern completion, symbol-digit substitution, tower rearranging and trail-making path tests. We also observed evidence of an observational association between being breastfed and an increase in years of schooling and a decrease in number of offspring (after controlling for covariates; Supplementary Table 11). In the case of the cardiometabolic diseases, being breastfed was associated (p < 0.05) with decreased risk of angina and heart attack, but not stroke or type-II diabetes after controlling for covariates (**Supplementary Table 12**). Being breastfed was associated with increased LDL, HDL and total cholesterol, and decreased triglycerides, BMI, and blood pressure (Supplementary Table 11). Finally, breastfeeding was observationally associated with increased risk of hayfever, rhinitis, and eczema (Supplementary Table 12).

**Figure 2:**
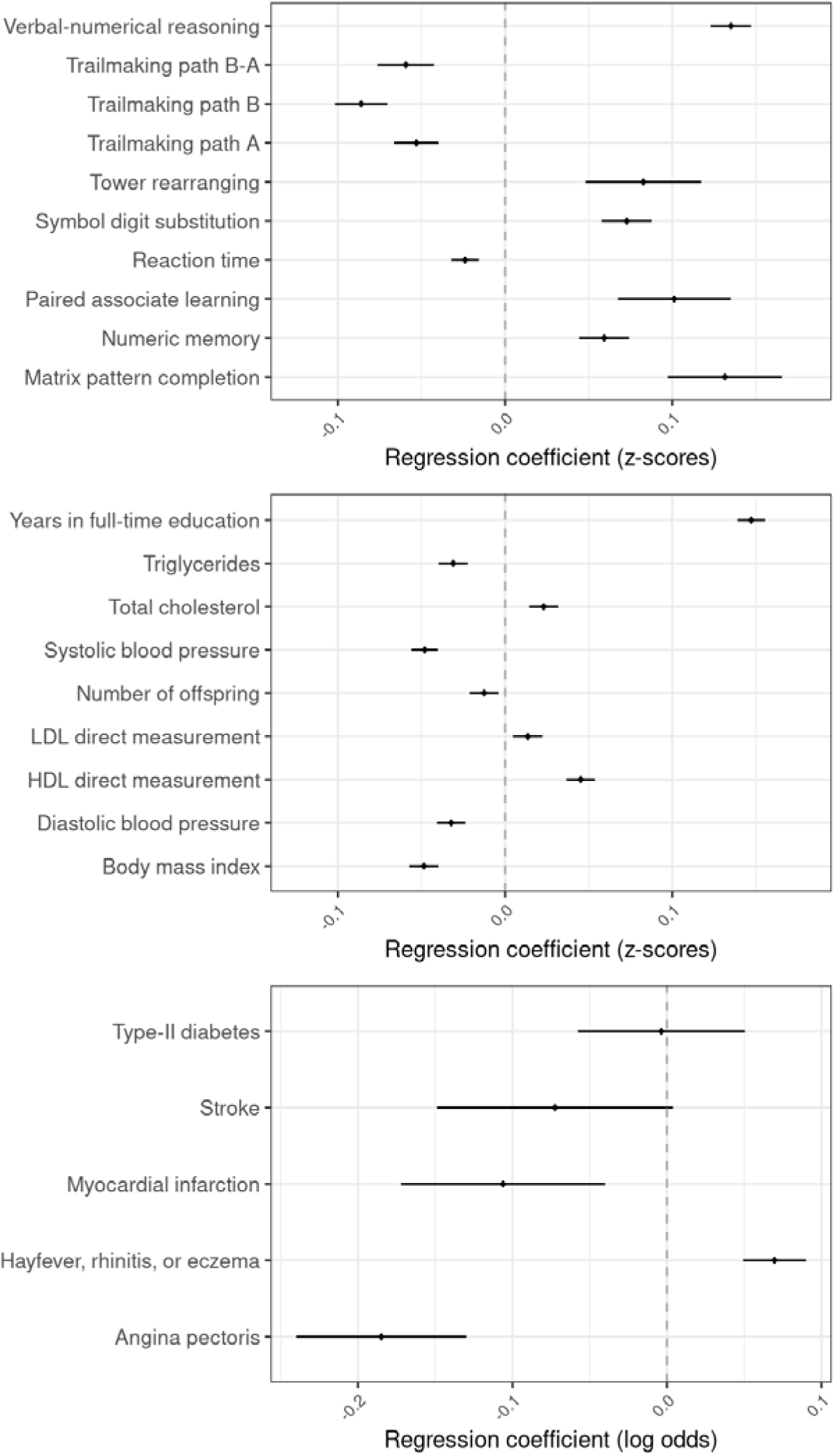
Association between breastfeeding and cognitive, noncognitive quantitative, and binary outcomes. Results are presented as change in z-score of the outcome (for quantitative traits) or change in log odds of being affected by the outcome when individuals are breastfed. All models were adjusted for age, sex, genetic principal components, and Townsend deprivation index. Error bars represent 95% confidence intervals.

### Association between FADS2 variants and fatty acid levels

We confirmed that all three *FADS* variants showed strong evidence of association with serum fatty acid levels in the expected directions (i.e., the mediating biomarkers through which breast feeding is thought to exert its beneficial effects; **Supplementary Table 2**) (49,53,82). All the variants were associated with DHA blood serum levels and with the ratio of DHA to total fatty acid levels. Alleles associated with an increase in DHA blood levels were associated with decreased LA blood serum levels and the ratio of LA to total fatty acids. All the DHA-increasing alleles were also associated with increased amount of PUFA, n-3, and n-6 fatty acids in the blood serum and their ratio to total fatty acids, and decreased n-6:n-3 ratio.

### FADS2 main effect and FADS2-by-breastfeeding interaction

All three *FADS2* variants were in strong LD with one another (*r* ^2^≥ 0. 68; Supplementary Table 1). We found little evidence for a main effect of rs174575 on verbal-numerical reasoning in the UK Biobank, when modelling rs174575 as recessive for the G allele as Caspi *et al.* (49) did (**Figure 3**). However, when modelling rs174575 under an additive genetic model (**Figure 4a**), additional copies of the DHA-increasing (C) allele were instead associated with *decreased* verbal-numerical reasoning (Figure 4, **Supplementary Table 13**).We found no evidence of interaction between rs174575 and breastfeeding on verbal-numerical reasoning, regardless of the covariate adjustment strategy (Supplementary Figure 1), both assuming a recessive model (Figure 3) or an additive model (Figure 4a and **Supplementary Figure 1**). The other SNPs showed similar results (**Supplementary Tables 13 to 16**). However, when extending the sample to non-White British participants, we observed evidence for a small interaction in verbal-numerical reasoning when the model did not account for PCs (**Supplementary Figure 4** and **Supplementary Table 17**).

**Figure 3:**
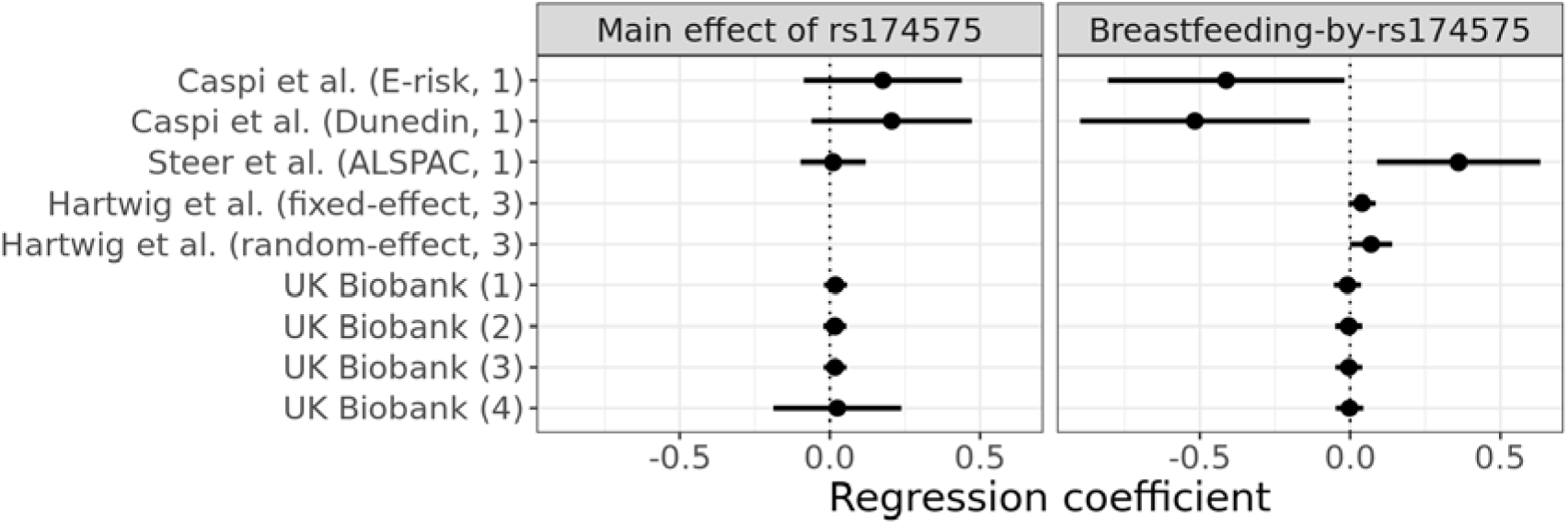
Effect estimates for the rs174575 main effect on verbal-numerical reasoning in the UK Biobank and in previous studies, and estimates of its interaction with breastfeeding. E-risk: Environmental risk study from Caspi *et al.* (49); Dunedin: Dunedin Multidisciplinary Health and Development Study from Caspi *et al.* (49); ALSPAC: Avon Longitudinal Study of Parents and Children from Steer *et al.* (50); UK Biobank: effect estimate derived by the present study on UK Biobank. (1) unadjusted; (2) adjusted for age and sex; (3) adjusted for age, sex, and principal components; (4) adjusted for age, sex, principal components, and pairwise interactions across covariates(81). Hartwig *et al.* (53) did not report an estimate of the main effect of rs174575. We derived the regression coefficients for Caspi *et al.* (49) from the mean and standard deviation of IQ for each subgroup reported in Table 1. The regression coefficients are expressed in change in standard deviations of Verbal-Numerical reasoning for change in genotype of rs174575 (modelled as a recessive effect of the C allele) for the main effect of rs174575, and in change of standard deviations of verbal-numerical reasoning for homozygous of the recessive alleles and breastfed individuals for the interaction term. Note that the large standard errors around the recessive main effect estimate in UK Biobank (4) are due to the covariate interaction terms correlating strongly with the recessive SNP coding. Error bars represent 95% confidence intervals.

**Figure 4:**
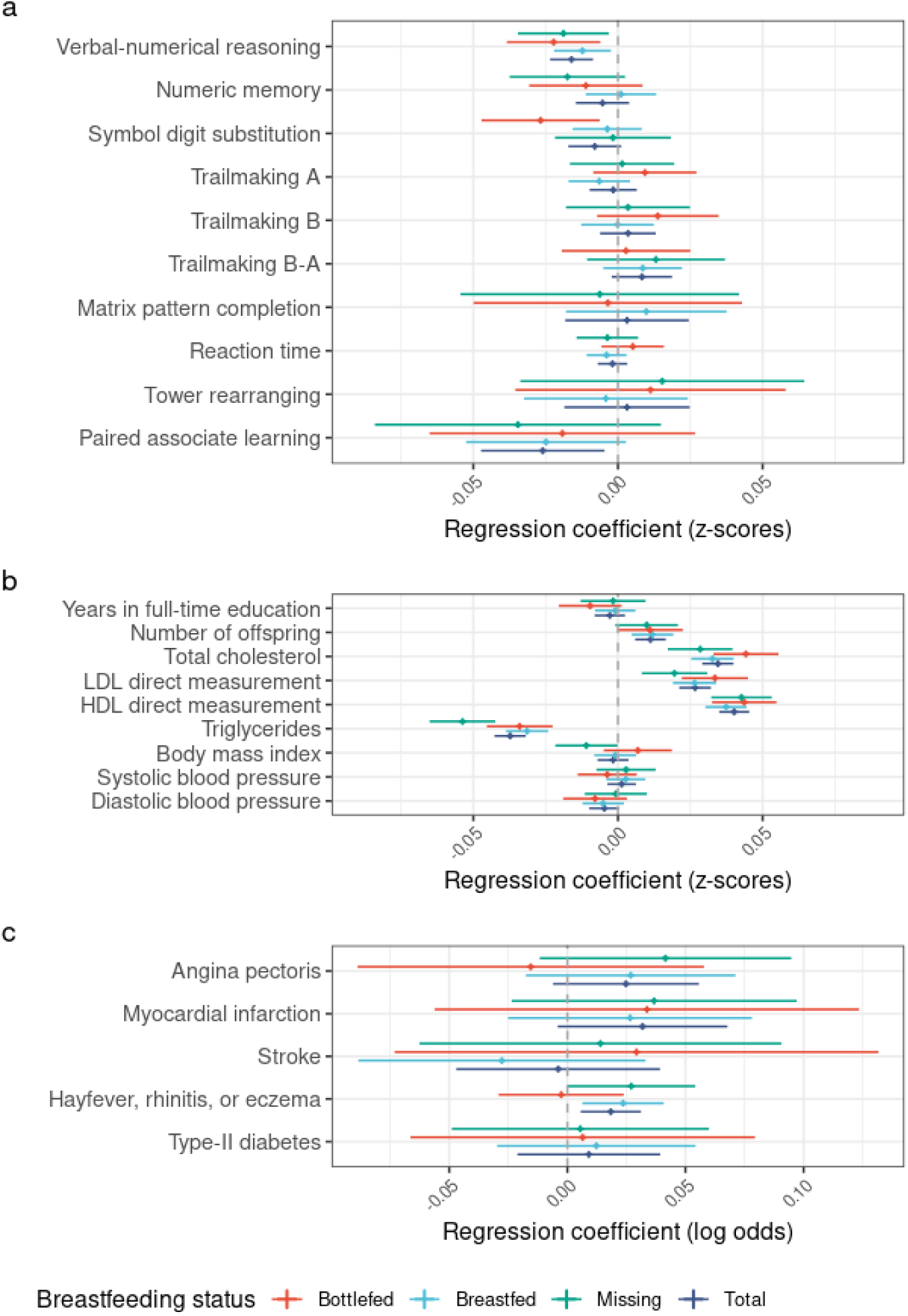
Results from the association analysis between rs174575 and all the outcomes by breastfeeding status, assuming an additive genetic effect of the DHA-increasing allele. The results are shown for rs174575 adjusting for age, sex, and principal components. a) results for the cognitive traits; b) results for the non-cognitive continuous traits; c) results for the binary traits (affected-unaffected). The results are expressed in change of standard deviations of the phenotype per increase in effect allele copies (for a and b) or change in natural log odds of being affected by the phenotype per increase in effect allele copies (for c). Error bars represent 95% confidence intervals.

The DHA increaser alleles at the three SNPs showed strong evidence of association with increased total cholesterol, LDL cholesterol, HDL cholesterol and number of offspring, and decreased triglycerides, BMI and blood pressure. Interestingly, assuming a recessive model at these same SNPs of the sort espoused by Caspi *et al*. (49), attenuated the evidence for association at most of the loci/phenotypes by several orders of magnitude. Similar, to the cognitive traits, the magnitude of the associations did not appear to be affected by breast-feeding status (**Supplementary Table 13 and 15**).

## Discussion

We failed to replicate an interaction between *FADS2* genotype and self-reported breastfeeding status on any of the measures of cognitive functioning we investigated, or any of the other traits we examined. Our study constitutes the largest attempt to replicate the Caspi *et al.* (49) interaction findings to date, and our results are similar to most, but not all previous attempts at replicating this interaction. A previous study by Steer *et al.* (50) also reported an interaction between breastfeeding and *FADS2* genotype on IQ in children from the Avon Longitudinal Study of Parents and Children (ALSPAC), a population-based longitudinal cohort with similar allele frequencies to E-risk and using the same family of IQ tests (Wechsler Intelligence Scale). However, the form of the reported interaction in Steer *et al.* (50) was different to that of Caspi *et al*. (49): Steer *et al.* (50) found a positive effect of breastfeeding on IQ in individuals homozygous for the G allele at the rs174575 variant, but not for carriers of the C allele. The two studies further differed in the way of reporting breastfeeding (i.e., prospectively in ALSPAC; retrospectively at age 2 or 3 in Dunedin and E-risk) and age at IQ testing (a combined measure of age 7, 9, 11, and 13 in Dunedin; 5 in E-risk; age 8 in ALSPAC).

An Australian twin study by Martin *et al*. (51) also failed to replicate the *FADS2-*by-breastfeeding interaction on IQ using a measure of cognitive functioning at age 16 and controlling for maternal SES. They also extended the analysis to rs174583, a non-palindromic SNP (T-C) in LD with the palindromic rs174575 (C-G), alongside the two previous variants, with similar results. Groen-Blokhuis *et al.* (52) investigated the same interaction for rs1535 ad 174575 on IQ measures at age 5, 7, 10, 12, and 18 (assessed with different scales), educational attainment, and maternally rated overactivity and attention problems in a different sample of twins from the Netherlands. Despite finding a small improvement in educational attainment, overactive behaviour, and IQ in breastfed twins compared to bottle-fed twins, they did not provide any evidence of an interaction between rs174575 and breastfeeding on childhood IQ or any of the other traits.

The largest and most comprehensive attempt at replicating the Caspi *et al*. (49) interaction was a study by Hartwig *et al.* (53) (12,077-13,202 individuals) who conducted a pre-registered (83) meta-analysis of studies, including the ALSPAC and Dunedin cohorts (49,50). The study excluded twin-based cohorts and limited the analysis to individuals of European ancestry, controlling for ancestry-informative PCs where possible, but failed to provide evidence of an interaction between breastfeeding and rs174575 on childhood IQ.

Most of these previous studies have been small, which historically has been typical of candidate gene studies (84–87). Our results seem to align with the findings of Hartwig and colleagues (53), which constitutes the best evidence prior to this study due to a rigorous pre-registered protocol, control for ancestry informative PCs where possible, and a large sample size (83). There could be several reasons why a gene-by-environment interaction was observed in some of the previous studies (viz., Caspi *et al.* (49) and Steer *et al.* (50)), but not others. A common criticism of candidate gene association studies is lack of sufficient control for population stratification (85,88). Failure to account for latent underlying differences in ancestry may induce spurious associations between traits and genetic variants that are functionally unrelated to the traits of interest(89). The Caspi *et al.* (49) and Steer *et al.* (50) studies attempted to control for population stratification by restricting their samples to a subset of homogeneous individuals, based on self-reported ancestry. However, this approach is often not sufficient to control for fine-scale genetic differences between populations and so it is likely that residual stratification was present. The use of ancestry-informative PCs as covariates is a common and effective way of controlling for population stratification in genetic association studies, as the PCs more accurately model individuals’ ancestry and capture more nuanced differences in latent population substructure beyond the major ancestral groups, thereby reducing the chance of spurious findings (90). In the present study, we accounted for population stratification in two ways: by using an ancestrally-homogeneous subset of UK Biobank individuals (as assessed by self-reported ancestry and scores on ancestrally-informative PCs (54)), and by fitting the PCs themselves as covariates.

Furthermore, when repeating the analysis for a larger, less ancestrally homogeneous sample of individuals (Supplementary Figure 4), we detected a weak genotype-by-breastfeeding interaction effect for rs1535 when not controlling for ancestry informative PCs-but this interaction disappeared after controlling for PCs. Our finding aligns with Hartwig *et al.* (53), where adjustment for PCs explained a substantial portion of the genotype-by-breastfeeding interaction in studies where this information was available. The implication is that previous positive findings may have been a result of inadequate control for population stratification, rather than genuine SNP-by-breastfeeding interaction effects.

Gene-by-environment interaction studies have also been criticised for not accounting for gene-by-covariate, environment-by-covariate, and covariate-by-covariate interactions. First suggested by Yzerbyt *et al.* (91) and highlighted by Keller (81) in the context of interaction studies with candidate variants, not accounting for pairwise interactions across covariates can bias the effect estimate for the interaction term. In the context of the present study, we performed the adjustment for the rs174575 variant, but we did not observe a substantial impact on the point estimates (which were close to the null). However, the standard error for the main effect estimate of the SNP drastically changed when fitting all the pairwise interactions across covariates (presumably because of high correlation between the recessive coding and the additional interaction terms in the model; Figure 3).

Finally, both Caspi *et al.* (49) and Steer *et al.* (50) fit different socioeconomic and physical covariates in their models, including SES (in both studies), maternal education, low birthweight, pre-term gestation, home environment, and parenting (in Steer *et al.* (50)). The use of heritable covariates to investigate genetic associations has the potential to bias the association or, in absence of a causal link, induce a spurious association, depending on the trait’s latent causal structure (92). Thus, it is possible that the interaction detected by the previous studies could have been partly or fully induced due to conditioning on a collider (93). In the context of the present study, we limited our covariate adjustments to age, sex, and PCs.

We provide some evidence for a main (additive) effect of rs1535, rs174575, and rs174583 on cognitive development, with all three SNPs showing an effect on the most commonly used proxy for cognitive development in UK Biobank (53,68,94–96), verbal-numerical reasoning. However, the direction of effect for verbal-numerical reasoning is opposite to what we would expect: since our analyses are aligned to the DHA-increasing allele, an increase in the effect allele count should lead to an increase in availability of DHA in breastmilk, and hypothetically an increase in cognitive ability. However, our results involve an inverse association.

To determine whether this discrepancy reflects a genuine association in the opposite direction, we compared our results with a previous GWAS meta-analysis on intelligence (96) (N = 269,867), which included unrelated participants from UK Biobank who also responded to the questionnaire on verbal-numerical reasoning (N = 195,653; also present in our study). In the case of a genuine genetic association, we would expect the increase in sample size to improve our evidence of an association and the direction of effect to align between the two samples. While the direction of effect for all three SNPs aligns with what we observed in the present study, the additional cohorts integrated in the meta-analysis improved the evidence of an association for only two SNPs (rs1535 and rs174575), while the p-value for rs174583 remained roughly unchanged ( *p=* 5. 66× 10^−5^ in Savage *et al.* (96), *p=* 4. 45× 10^−5^ in the present study; Supplementary Table 21). Due to the increase in sample size in the meta-analysis (N = 74,214) and the strong LD with rs1535 (*r* ^2^= 0. 96, we would expect rs174583 to benefit from the inclusion of additional cohorts, hinting at the possibility of a false positive in the UK Biobank.

Another possibility is that the SNP in *FADS2* are capturing an independent signal from a causal variant within. When looking at the LD structure surrounding *FADS2,* the SNPs appear to be independent from the nearest genome-wide significant top hit (rs77128898), making it unlikely they are tagging signals from that region. On the other hand, as the region immediately surrounding the variants in *FADS2* hosts several other genes, the SNPs in *FADS2* may still be tagging an independent signal not originating from *FADS2*, namely: *MYRF*, a transcription factor involved in myelination of the central nervous system; *FADS1,* another desaturase part of the DHA and AA biosynthesis; *MIR611* and *FEN1*, involved in the alteration of miRNA and Okazaki fragments, respectively; and *TMEM258*, involved in protein N-linked glycosylation (**Supplementary Figure 5**). Nevertheless, inference on the causal mechanisms for the region is a difficult process that require inferences on the causal variants (often involving fine-mapping (97), a statistical procedure attempting to determine the source of signal within a genome-wide significant region) and strong evidence of the association of the trait with the region. Further investigation of the region surrounding *FADS2* is required to elucidate its role in cognitive development.

Our results from the UK Biobank align with much of the current literature suggesting a positive effect of breastfeeding on cognitive traits, after controlling for covariates (8). Nevertheless, our analyses are based upon observational data. Observational association studies of breastfeeding are typically affected by residual confounding, selection bias, and reverse causation, as has been noted in previous studies involving IQ (7,76,98) (where maternal IQ and socioeconomic status may influence the likelihood of being breastfed, or reporting to be breastfed) and atopy (99–101) (where early signs of atopic allergy may cause the offspring to be breastfed for longer). While some quasi-experimental designs robust to reverse causation have also observed a positive effect of breastfeeding on IQ (10,102), the breastfeeding literature is likely subject to a considerable publication bias (103), inflating the ratio of reported positive to null findings. Ultimately, more studies are necessary to dissect the causal relationship between breastfeeding and cognition.

Several limitations of the present study arise from the use of self-reported breastfeeding status. Recollection of their own breastfeeding status by the UK Biobank participants is susceptible to self-report bias, as the recall is likely dependent on the information being passed on by their mother and/or other relatives. The UK Biobank participants’ year of birth (1934—1971) corresponds to a period generally regarded as having low breastfeeding initiation and cultural taboos related to breastfeeding practices in the UK (104). While systematic collection of data on breastfeeding practices succeeded that time (the Infant Feeding Survey, the longest-running survey on infant feeding practices in the country, started only in 1975 (105)), data from other countries generally show a steady decline in breastfeeding from 1930s to 1970s, when infant formula became the prevalent way of feeding infants (106), and indeed the UK retains a low uptake of breastfeeding to this day (107).Conversely, around ¾ of the UK Biobank participants with data on the question reported being breastfed as infants, hinting at potential issues with the self-reported measure of breastfeeding available in the UK Biobank. Due to the promotion of breastfeeding as the “healthy” way of feeding infants, together with a sampling skewed more towards healthier, older, and female individuals (108) (i.e., more likely to have been exposed to breastfeeding promotion campaigns), it is possible that the measure could suffer from non-differential misclassification bias (109): participants would be more likely to have answered they were breastfed than bottle fed, when in doubt. As we expect these variants to show an effect in the breastfed individuals, this could bias our estimates towards the null.

Other potential issues that could impact our estimates are due to the varying duration of breastfeeding across participants. Early weaning well before the currently recommended six months of exclusive breastfeeding (1) has been reported to be common practice before 1970 (110), and it is still occurring in the UK to this day (111). As we expect a change in the concentration of DHA in breastmilk according to the levels of *FADS2* genotype only within the breastfed group, if an interaction were present, a shorter breastfeeding duration would attenuate the effect of DHA on the outcome.

The quality of cognitive measures in the UK Biobank presents further challenges to our findings. The previous literature describing the effect of breast feeding on IQ is heterogeneous in terms of scales used for IQ assessments and age at assessment, but it is predominantly composed of validated scales of child IQ. In contrast, the cognitive measures available in the UK Biobank were taken in mid-to late adulthood and used as a proxy for early childhood cognitive development. While absolute cognitive performance declines after early adulthood (112,113), intelligence does show rank-order stability over time (114,115) (i.e., an individual’s position according to a reference population), which is the rationale behind the widespread use of standardised IQ scores. Thus, by assuming temporal stability and accounting for age and cohort differences (116), adult cognitive performance can be used as a proxy for childhood intelligence to observe differences across subgroups, although with different temporal stability across ranks (117,118).

Despite being mostly based on established psychometric measurements, the adoption of non-validated tests without data on their standardisation poses difficulties in the interpretation of our results. In particular, it is difficult to compare the UK Biobank participants’ performance with the general population. As the effect of a supposed breastfeeding-by-*FADS2* interaction might not be uniform across the whole population, the lack of standardisation prevents inferences drawn from quantile-based subgroup analyses. Further, the relative homogeneity in stability of the UK Biobank cognitive tests (116), together with the known sampling biases(108), suggests the lower ranks of cognitive abilities might be underrepresented, which might be also affected differently by the gene-by-breastfeeding interaction.

Finally, some studies (70,119) have noted that the most-commonly used measure used to proxy intelligence in the UK Biobank (labelled *fluid intelligence* by UK Biobank, here and in the literature often named *verbal-numerical reasoning* (68,94,95,119)) might capture a blend of crystallised and fluid abilities. Following Hebb and Cattel’s paradigm on intelligence (120,121), fluid intelligence represents what is commonly known as “intelligence”, a task-independent cognitive performance, while crystallised abilities reflect skills acquired through learning and practice over time (i.e., developed through the investment of fluid intelligence in new tasks). This distinction can be also observed in the underlying biology of the trait, with fluid and crystallised skills showing different genetic associations and pathways (122); verbal (crystallised) and non-verbal (fluid) skills have been also observed to have different topological correlates during brain development (123). As the *FADS2-*by-breastfeeding interaction we have investigated should influence predominantly fluid skills, with a lesser impact on crystallised skills in later life, capturing crystallised abilities could attenuate the effect of a possible interaction, reducing power to detect an effect with our current sample size.

Despite benefitting from the largest sample size to date, our study does not provide evidence for a *FADS2*-by-breastfeeding interaction on any of the traits we investigated, after strict control for population structure. Our findings align with smaller studies that have also failed to detect the interaction after controlling for population structure through PCs (53). We provide evidence for a small main effect of the *FADS2* variants (rs1535, rs174575, rs174583) on a subset of our traits (viz., the verbal-numerical reasoning scale for cognitive traits, reproductive success, and lipid measures).

## Supporting information

STREGA checklist

## Data Availability

The UK Biobank data described in the manuscript is available to all bona fide research upon application and approval. Code book and analytic code are available online at https://github.com/GiulioCentorame/FADS-by-breastfeeding/

## Acknowledgements

We would like to thank the UK Biobank participants, interviewers, nurses, and staff that made this study possible. We wish to acknowledge The University of Queensland’s Research Computing Centre (RCC) for its support in this research.

## Statement of authors’ contribution to manuscript

GC, NW, GH, GDS, and DME designed the research; GC analysed the data; GC, NW, and DME wrote the paper. GW provided help on the analysis for the blood pressure phenotypes. DME had primary responsibility for final content. All authors have read and approved the final manuscript.

---

**Supplementary Figure 1:**
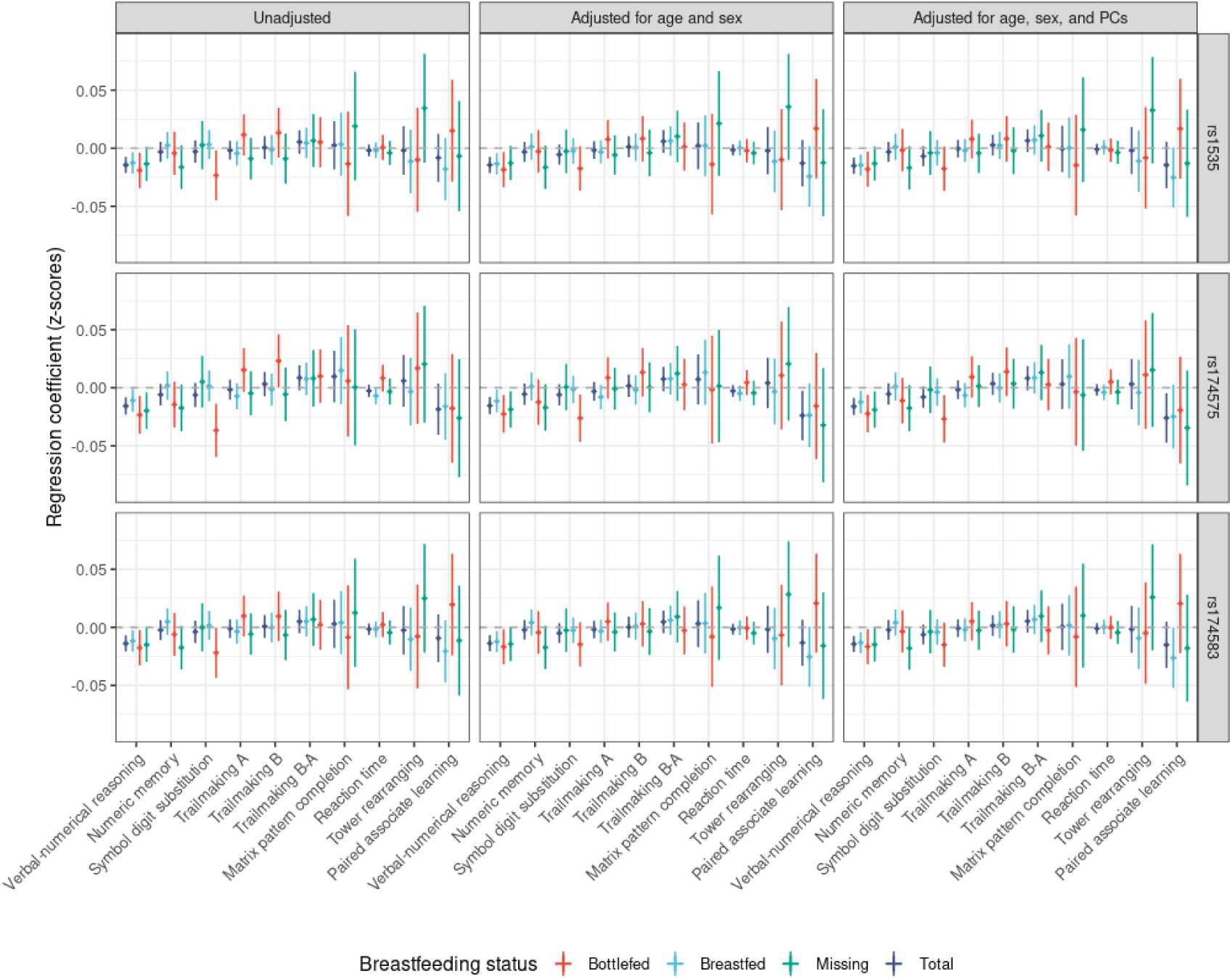
Results from the association analysis between the three FADS2 SNPs (additive effect) and cognitive traits stratified by breastfeeding status. The results are expressed in change of standard deviations of the phenotype per increase in effect allele copies in each SNP. Error bars represent 95% confidence intervals.

**Supplementary Figure 2.**
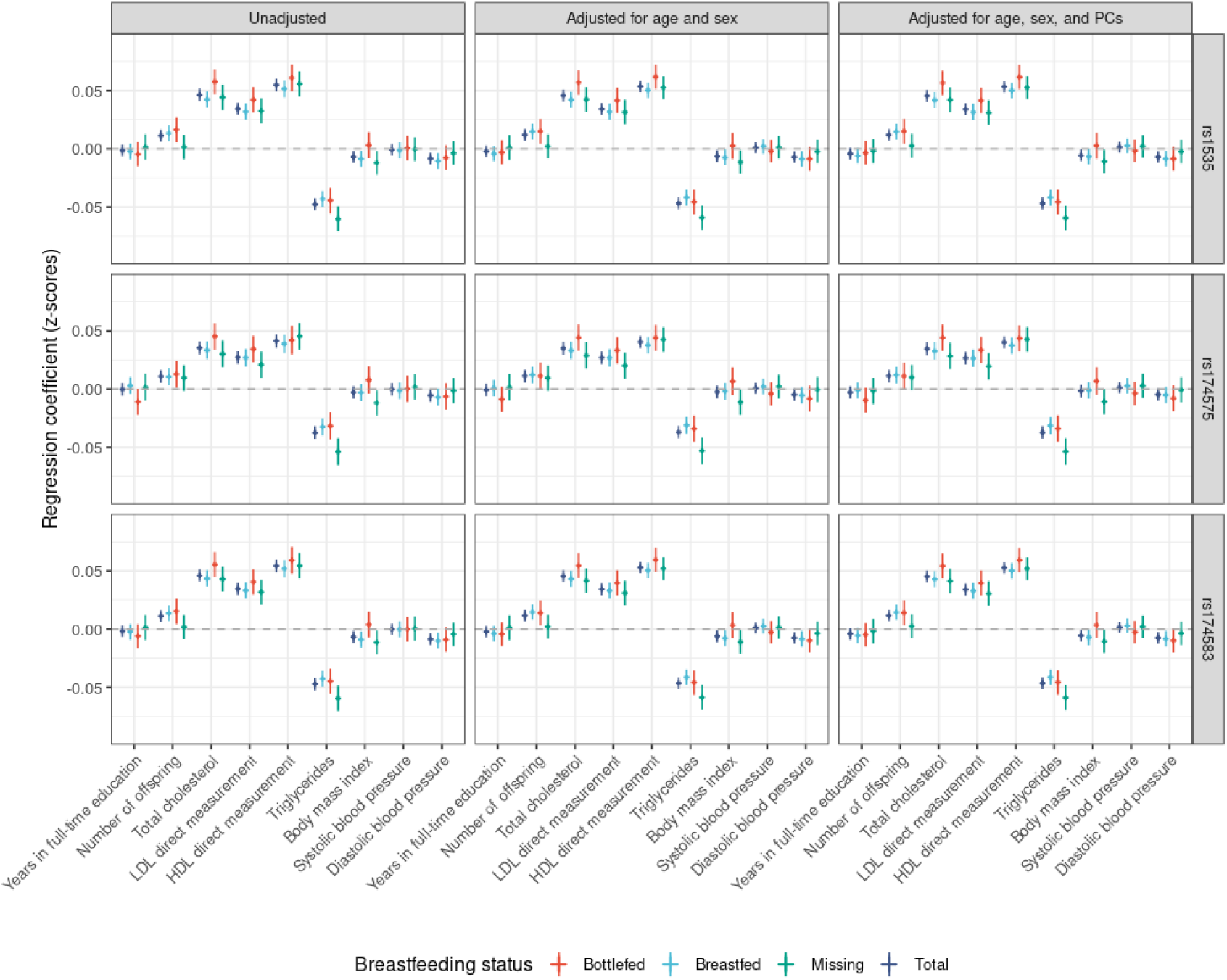
Results from the association analysis between the three FADS2 SNPs (additive effect) and noncognitive continuous traits stratified by breastfeeding status for all adjustment strategies. The results are expressed in change of standard deviations of the phenotype per increase in effect allele copies in each SNP. Error bars represent 95% confidence intervals.

**Supplementary Figure 3:**
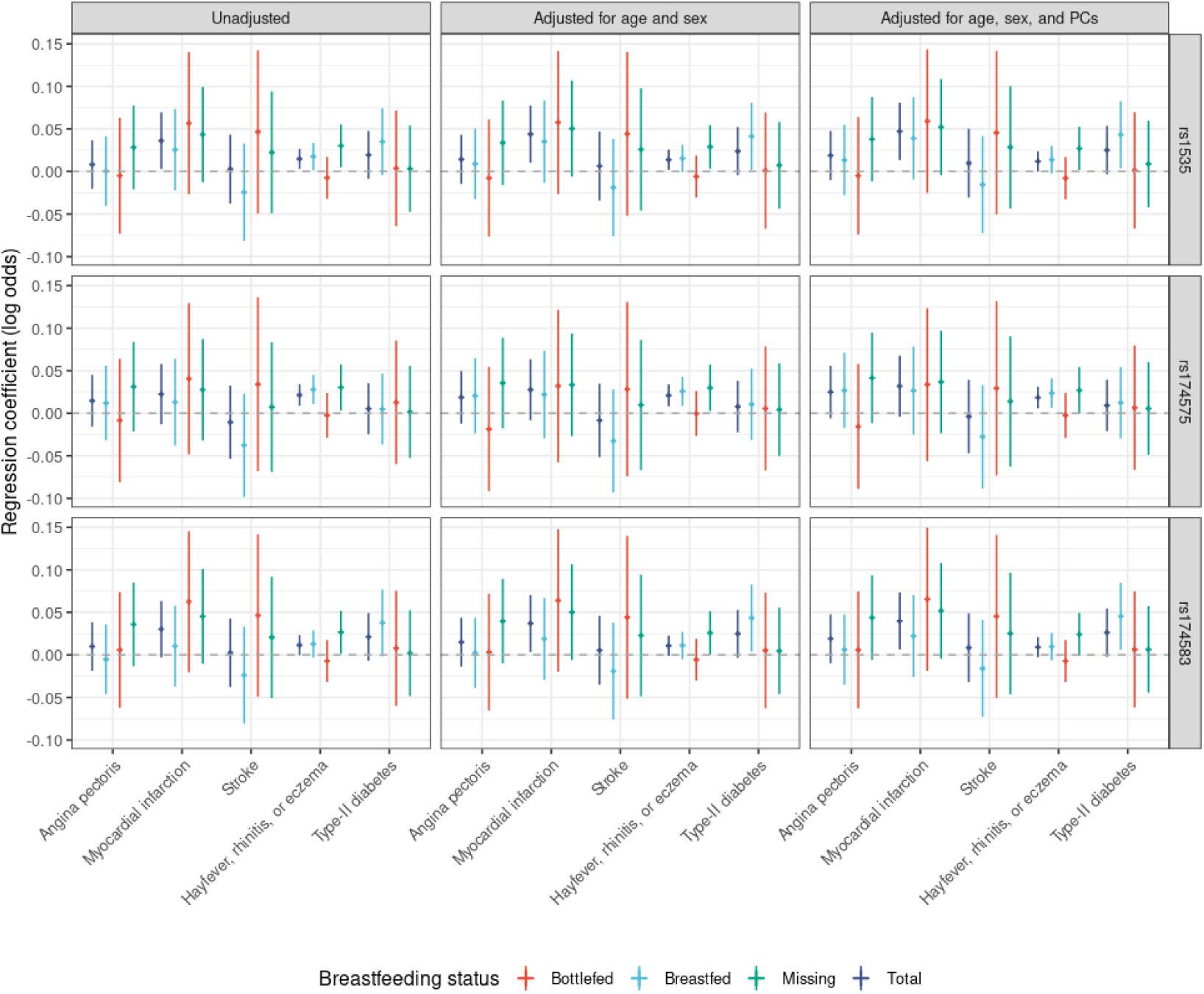
Results from the association analysis between the three FADS2 SNPs (additive effect) and binary traits (affected-unaffected) stratified by breastfeeding status for all adjustment strategies. The results are expressed in change of natural log odds of being affected by the phenotype per increase in effect allele copies in each SNP. Error bars represent 95% confidence intervals.

**Supplementary Figure 4:**
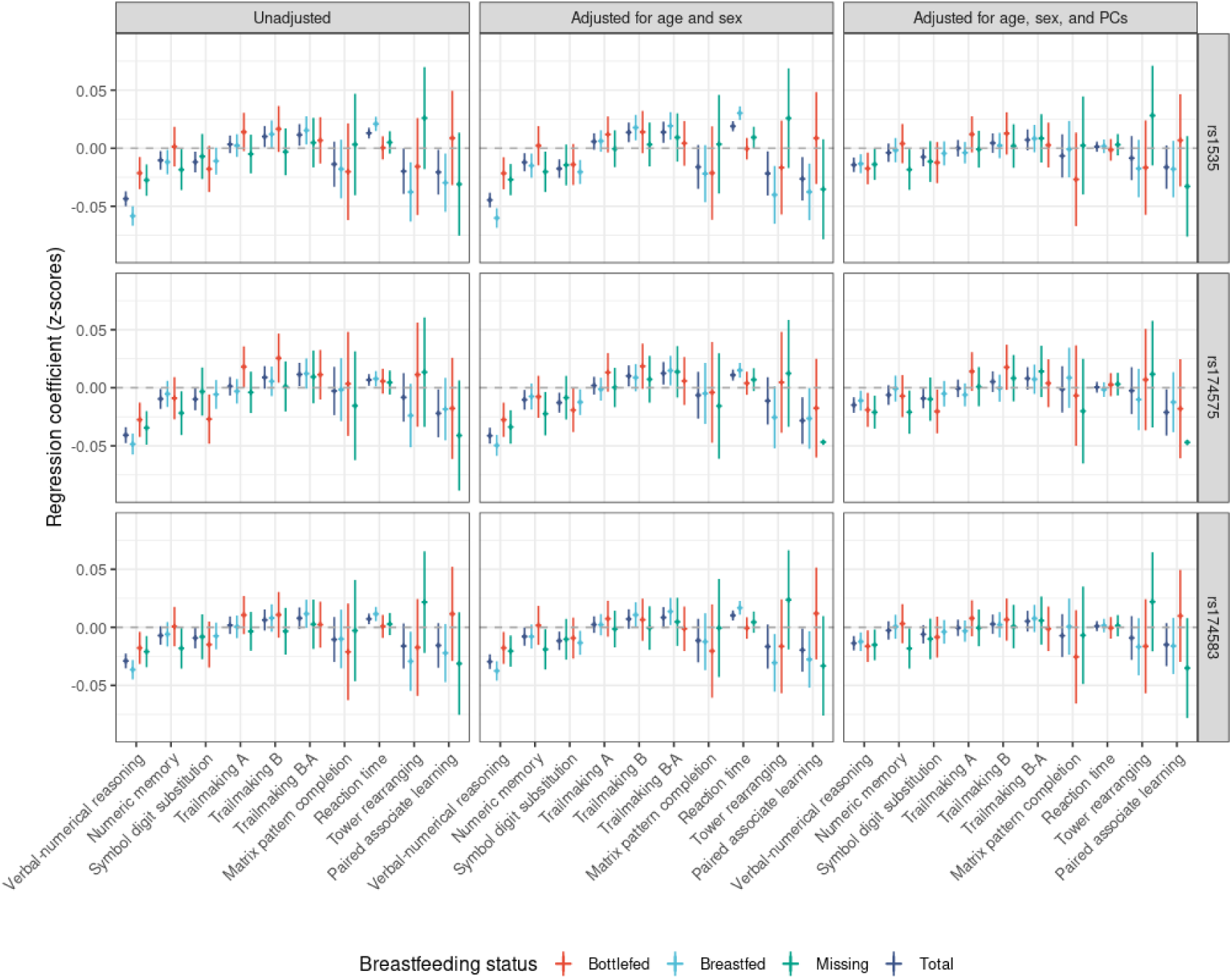
Results from the association analysis between the three FADS2 SNPs (additive effect) and cognitive traits stratified by breastfeeding status for all adjustment strategies, all ancestries. The results are expressed in change of standard deviations of the phenotype per increase in effect allele copies. Error bars represent 95% confidence intervals.

**Supplementary Figure 5:**
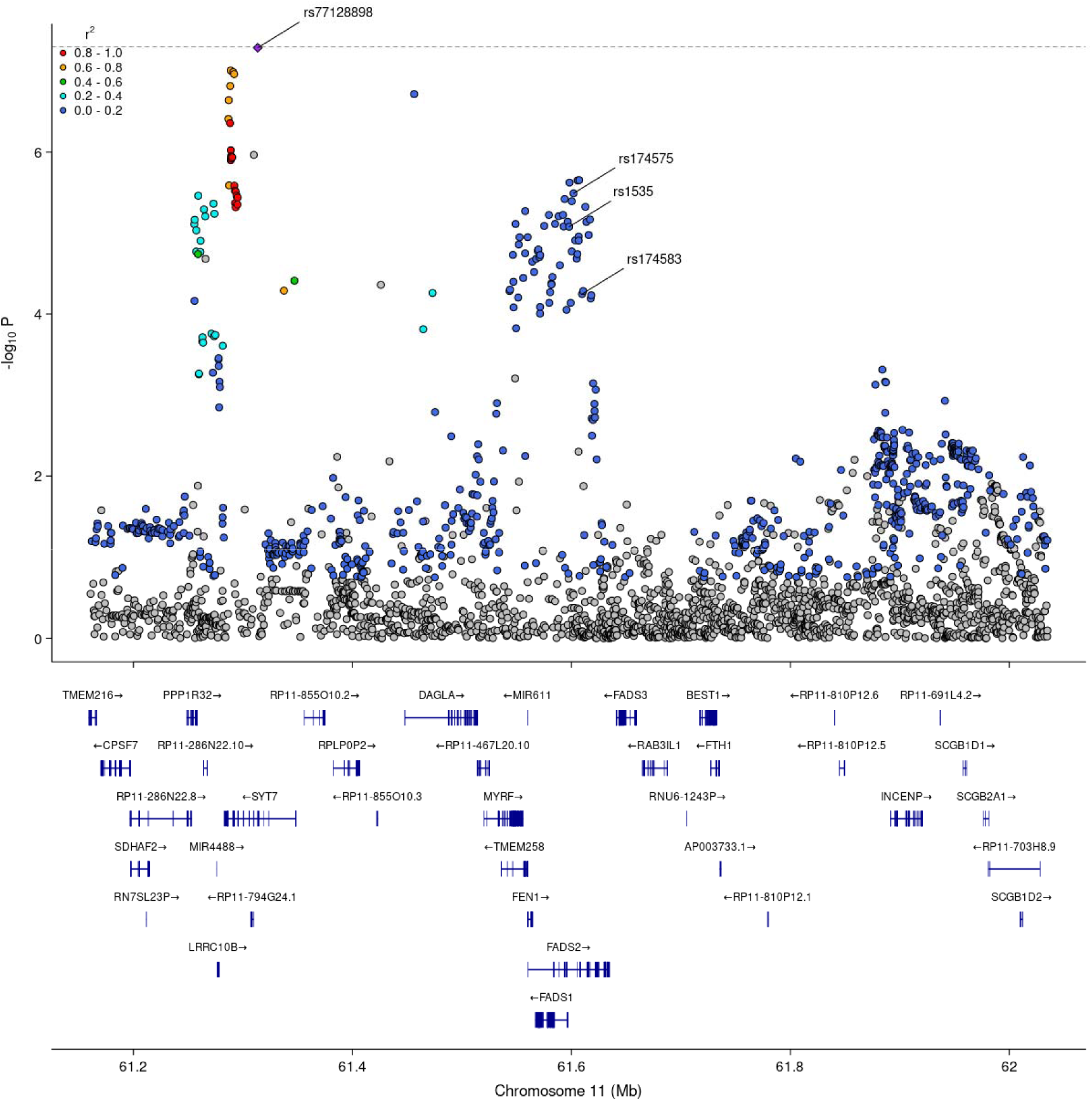
local plot of LD structure in the region surrounding FADS2. Data from Savage *et al.* (96). *r*^2^ is relative to the nearest top hit (rs77128898) based on the CEU panel (Northern Europeans from Utah) from 1000 Genomes (124). Gene tracks were obtained from Ensembl release 75 (125). The dashed line represents the genome-wide significance threshold ( *p=* 5× 10^−8^).

